# Urine lipoarabinomannan concentrations among HIV-uninfected adults with pulmonary or extrapulmonary tuberculosis disease in Vietnam

**DOI:** 10.1101/2023.07.17.23292752

**Authors:** Nguyen B. Hoa, Mark Fajans, Hung Nguyen Van, Bao Vu Ngoc, Nhung Nguyen Viet, Hoa Nguyen Thi, Lien Tran Thi Huong, Dung Tran Minh, Cuong Nguyen Kim, Trinh Ha Thi Tuyet, Tri Nguyen Huu, Diep Bui Ngoc, Hai Nguyen Viet, An Tran Khanh, Lorraine Lillis, Marcos Perez, Katherine K. Thomas, Roger B. Peck, Jason L. Cantera, Eileen Murphy, Olivia R. Halas, Helen L. Storey, Abraham Pinter, Morten Ruhwald, Paul K. Drain, David S. Boyle

## Abstract

Lipoarabinomannan (LAM) is a Mycobacterial cell wall glycolipid excreted in urine, and a target biomarker of rapid diagnostic tests (RDTs) for tuberculosis (TB) disease. Urine LAM (uLAM) testing by RDT has been approved for people living with HIV, but there is limited data regarding uLAM levels in HIV-negative adults with TB disease. We conducted a clinical study of adults presenting with TB-related symptoms at the National Lung Hospital in Hanoi, Vietnam. The uLAM concentrations were measured using electrochemiluminescent immunoassays and compared to a microbiological reference standard (MRS) of sputum, GeneXpert Ultra and TB culture. Additional microbiological testing was conducted for possible extrapulmonary TB, when clinically indicated. Among 745 participants enrolled, 335 (44.9 %) participants recruited from the pulmonary TB wards (PR-PTBW) and 6 (11.3%) participants recruited from the EPTB wards (PR-EPTBW) had confirmed TB disease. The MRS positive cohort measured median uLAM concentration for S4-20/A194-01 (S/A) were 14.5 pg/mL and 51.5 pg/mL, respectively. The FIND28/A194-01 (F/A) antibody pair overall and TB-positive cohort measured mean uLAM was 44.4 pg/mL and 78.1 pg/mL, respectively. Overall, the S/A antibody pair had a sensitivity of 39% (95% Confidence Interval [CI] 0.33, 0.44) and specificity of 97% (95% CI 0.96, 0.99) against the MRS. The F/A antibody pair had a sensitivity of 41% (95% CI 0.35, 0.47) and specificity of 79% (95% CI 0.75, 0.84). The areas under the receiver operating curves were 0.748 for S/A and 0.629 for F/A. There was little difference between the S/A median uLAM concentration with pulmonary (55 pg/mL) and extrapulmonary (36 pg/mL) TB disease. With F/A the medians for pulmonary and extrapulmonary TB disease were 79% and 76.5% respectively. Among HIV-negative adults in Vietnam, concentrations of uLAM remained relatively low for people with TB disease, which may present challenges for developing a more sensitive rapid uLAM test.

## Introduction

Tuberculosis (TB) remains one of the leading infectious causes of death worldwide. Most TB infections occur among HIV-negative persons, but people living with HIV (PLWH) have a significantly higher individual risk of TB disease and case fatality rate [1,2]. In 2022, approximately 40% of people with TB disease were either not diagnosed or not reported to national disease surveillance systems [2]. Many people in TB-endemic settings may have limited access to care, and improved access to TB diagnostics could improve treatment initiation and reduce mortality [3].

Lipoarabinomannan (LAM) is a pathogen biomarker that has clinical utility for diagnosing TB disease [4]. LAM is a glycolipid in the Mycobacterial cell that is also secreted in exosomes and subsequently excreted in urine [5,6]. Capped derivatives of LAM are TB specific and detectable in urine [7–10]. The Alere Determine™ TB LAM Ag (Determine, Abbott Diagnostics, USA) is a rapid diagnostic test (RDT) approved by the World Health Organization (WHO) for use among PLWH with immune suppression and advanced disease [11–13]. However, this RDT is only approved for people with TB-related symptoms who have a CD4 <200 cells/mm^3^ [14]. As such, the only WHO-approved uLAM RDT has insufficient sensitivity for HIV-uninfected people with TB disease.

Currently, there is limited data on the concentration of urine LAM (uLAM) in HIV-uninfected adults with TB disease. Prior studies of uLAM concentrations used material predominantly from mycobacterial culture. which differs structurally from *in vivo* isolated LAM [15,16]. Most clinical validation studies using RDTs for uLAM have focused on PLWH and evaluated performance against microbiological reference standards (MRS, culture and/or GeneXpert) [17,18]. One study described two small cohorts from Peru and South Africa showed mixed results, and these were underpowered for HIV-uninfected participants [19]. A scaled clinical evaluation of uLAM using an ultra-sensitive LAM immunoassay has not been performed in HIV-uninfected adults. Our objective was to accurately quantify the concentration of uLAM and evaluate clinical diagnostic accuracy among HIV-uninfected adults presenting with TB-related symptoms using highly sensitive electrochemiluminescent (ECL) immunoassays [20]. We included adults with both pulmonary and extrapulmonary TB in a high TB-burden, low HIV setting of Hanoi, Vietnam.

## Materials and Methods

### Study Design and Participants

We conducted a prospective longitudinal cohort study of adults 18 years of age or older who had TB-related symptoms and presented for routine care at the National Lung Hospital (Hanoi, Vietnam) between October 2021 and April 2022. Presumptive participants recruited from the pulmonary TB ward (PR-PTBW) were recruited from both outpatient clinic and hospitalized respiratory inpatient ward.

Presumptive participants recruited from the extra pulmonary TB (PR-EPTBW) ward were recruited from an EPTB inpatient ward exclusively. Exclusion criteria included having received isoniazid preventive therapy within the previous 3 months, having received anti-TB treatment for more than 24 hours, or having a confirmed TB diagnosis at time of recruitment. Details of study recruitment, eligibility and exclusion criteria, and enrollment procedures are described in the supplementary materials. Eligibility criteria were developed to be representative of people who may be evaluated for pulmonary or extrapulmonary TB disease in Vietnam (see S1 Doc and S2 Doc).

### Ethical approvals

The institutional review boards (IRBs) at the National Lung Hospital (NLH, Hanoi, Vietnam; reference No: 26/21/CN-HDDD) and the Vietnam Ministry of Health (Hanoi, Vietnam; reference No: 95/CN-HDDD) approved the study and the PATH Office of Research Affairs (ORA) gave approval contingent on the NLH IRB decision. Written informed consent was obtained from each eligible participant prior to enrollment. The immunoassay testing at the PATH laboratory was determined as non-human subjects research by the PATH ORA before any analysis was performed on the samples.

### Study Procedures and Clinical Care

After enrollment, we collected sociodemographic and clinical history, and participants received a clinical evaluation as part of the routine standard of care. If a participant did not have a documented HIV test within the prior 6 months, then they were offered testing. HIV testing was offered using Determine HIV-1/2 (Abbot Diagnostics, Scarborough, ME) or Quick Test HIV 1 & 2 (Amvi Biotech, Ho Chi Minh City, Vietnam), and positive results confirmed with a second rapid test and finally an HIV ELISA (Murex HIV Ag/Ab combination, DiaSorin, Saluggia, Italy). For PR-PTBW, we collected separate sputum samples for Xpert MTB/RIF Ultra and MGIT liquid culture testing for all participants. For PR-EPTBW), we collected non-respiratory samples (CSF, pleural fluid, etc.) for MGIT testing and Xpert MTB/RIF Ultra testing, as clinically indicated. We obtained urine samples, which were frozen at –80 °C immediately after collection and until the lab-based testing. At 2 and 6 months after enrollment, we conducted follow-up phone calls to participants to evaluate clinical status and response to TB treatment, and searched clinical chart and laboratory records to ascertain incident diagnoses, repeat hospitalization, or treatment initiation. Approximately 30 mLs of urine from each participant was collected for analysis with the ECL immunoassays and the remaining material was retained to create a biorepository at PATH for future access to developers of uLAM diagnostic products.

### Evaluation of the clinical urine specimens using immunoassays

The concentration of uLAM in the urine specimens was measured using highly sensitive electrochemiluminescent (ECL) immunoassays [20]. We separately evaluated two different monoclonal capture antibodies, FIND28 (Foundation for Innovative New Diagnostics, Geneva, [20]) and S4-20 (Otsuka Pharmaceuticals, Tokyo, Japan [21]), that were biotinylated (EZ-Link Sulfo-NHS-LC-Biotinylation Kit, ThermoFisher Scientific, Waltham, MA, USA). For both capture antibodies, we used the same recombinant detector antibody, A194-01 (Rutgers University [9]), which was labeled with the GOLD SULFO-TAG NHS-Ester (Mesoscale Diagnostics [MSD], Rockville, MD, USA). Unbound labels (biotin or SULFO-TAG) were removed using desalting columns (40 kDa MWCO Zeba Spin, Thermo).

The biotinylated capture antibodies were coupled to U-PLEX plates (MSD) via biotin-streptavidin binding to U-PLEX linkers (MSD). Both antibody-linker conjugates were mixed with U-PLEX stop buffer (MSD) at a concentration of 0.29 µg/mL and 50 µL added to each well. Plates were incubated for 1 hour with shaking at 500 rpm to allow for the antibodies to self-assemble to their discrete and complimentary linker-binding sites to create a 2-plex immunoarray. Plates were washed 3X with 300 µL/well of 1X phosphate buffered saline + 0.05% Tween 20 (PBS-T, pH 7.5) using a ELX405R microplate washer (BioTek Instruments Inc., Winooski, VT).

The analysis of uLAM was performed by adding 25 µL of Buffer 22 (MSD) to each well followed by 25 µL of unconcentrated urine sample, standard, or control [20]. Plates were incubated at room temperature with shaking for 1 hour at 500 rpm to allow binding of uLAM to the capture antibodies. Plates were washed 3X with PBS-T and then 25 µL of SULFO-TAG detection antibody (2 µg/mL) in Diluent 3 (MSD) was added to each well with shaking for 1 hour. Plates were washed 3X with PBS-T and filled with 150 µL of 2X read buffer T (MSD) per well. The plates were read in a MESO QuickPlex SQ 120 plate reader (MSD) and the ECL from each individual array spot were measured and analyzed using the Discovery Workbench v4 software (MSD). The LAM used for the standard curve was derived from the *in vitro* culture of the TB strain Aoyama B (Nacalai USA, Inc., San Diego, CA) was dissolved in 1% BSA in water (w/v). Duplicate seven-point serial dilutions of the TB LAM stock (40,000 pg/ml to 2.44 pg/mL) in 2% BSA/1X PBS with a negative control were used to generate calibration curves.

The relationship of ECL to LAM concentration was fitted to a four-parameter logistic (4-PL) function. The uLAM concentration in each specimen was then calculated for both antibody pairs by back-fitting the ECL data to the 4-PL fit using proprietary software from MSD. A specimen was considered detected when the ECL signal of the sample correlated with signals above the limit of detection in the assay (LOD) determined from the software. The LOD was calculated by the software, based on signal generated by the calibrators and negative control and deviated slightly from plate to plate (S1 Table). As such we used the LOD established from each plate to score the test results and so afford greater accuracy, as opposed to earlier approaches of applying universal cutoff values [19,20]. Anything below the LOD was considered not detected. In cases where samples containing uLAM was above the upper limit of detection, then the sample was diluted in 1X PBS and reassessed.

### Reference Standards

The microbiological reference standard (MRS) was defined as having a positive result by either Xpert Ultra or Mycobacterial culture by MGIT. For PR-PTBW, a positive microbiological test result using a respiratory (sputum) specimen was required, while both a negative Xpert Ultra and MGIT culture result using a respiratory specimen was required to be considered negative. Two separate sputum samples were collected consecutively for Xpert Ultra and MGIT testing. The MRS for EPTB included tissue samples from pleura, lymph nodes, abdomen, or meninges. Due to the difficulty in using non-respiratory specimen for Xpert testing, when no Xpert testing was done for non-respiratory specimen, a negative EPTB cases would consist of a negative culture using a non-respiratory specimen. The clinical reference standard (CRS) was defined as either meeting the MRS definition or receiving empiric TB treatment (+/- compatible chest X-ray) within 2 months of enrollment at the discretion of the treating clinicians.

### Statistical Analyses

We report diagnostic accuracy of the ECL immunoassays via sensitivity, specificity, positive and negative predictive values, and positive and negative likelihood ratios, along with 95% confidence intervals estimated using the binomial exact (Clopper-Pearson) method. The primary reference standard was the MRS, with Xpert result alone, culture result alone and CRS reference standards presented for additional information. Comparison of sensitivities or specificities between the PR-PTBW and PR-EPTBW groups were made using Fisher’s exact test. Comparisons of sensitivities or specificities between mAb pairs within the same TB group were made using McNemar test. Receiver operating characteristic (ROC) curves were calculated to evaluate the overall discriminatory ability of each antibody (Ab) pair, and statistically compared using paired or unpaired ROC comparisons (R package pROC, v1.18), as appropriate [22]. We conducted all analyses using R version 4.1.2 [23]. All of the data generated in this study can be publicly accessed at Dataverse (https://doi.org/10.7910/DVN/AOL0LP).

## Results

We enrolled 780 participants from the wards (700 people with presumed PTB [PR-PTBW] and 80 people with presumed EPTB PR-EPTBW]). Of these, 11 participants were excluded due to the urine collection not being within 48 hours of enrollment, and 3 were excluded due to being HIV-positive, leaving 766 participants being evaluated by the MRS (693 PR-PTBW, 73 PR-EPTBW; Figure 1). From the PR-PTBW group, 1 participant could not be validated due to culture being available only on an extrapulmonary specimen (pleural fluid). From the PR-EPTBW group, 20 participants could not be validated because non-pulmonary specimens were not available for validation testing. Of these, 15 had only a pulmonary sample collection for validation, one had only respiratory (sputum) specimen collected for Xpert testing and none for MGIT testing, and four had no sample collected for Xpert or MGIT testing. The reasons for missing samples on these four were that no sample fluids were available for collection from cases of presumed spinal TB, presumed intestinal TB and presumed bone TB respectively. A total of 692 PR=PTBW and 53 PR-EPTBW participants were included in our analyses.

**Figure 1.**
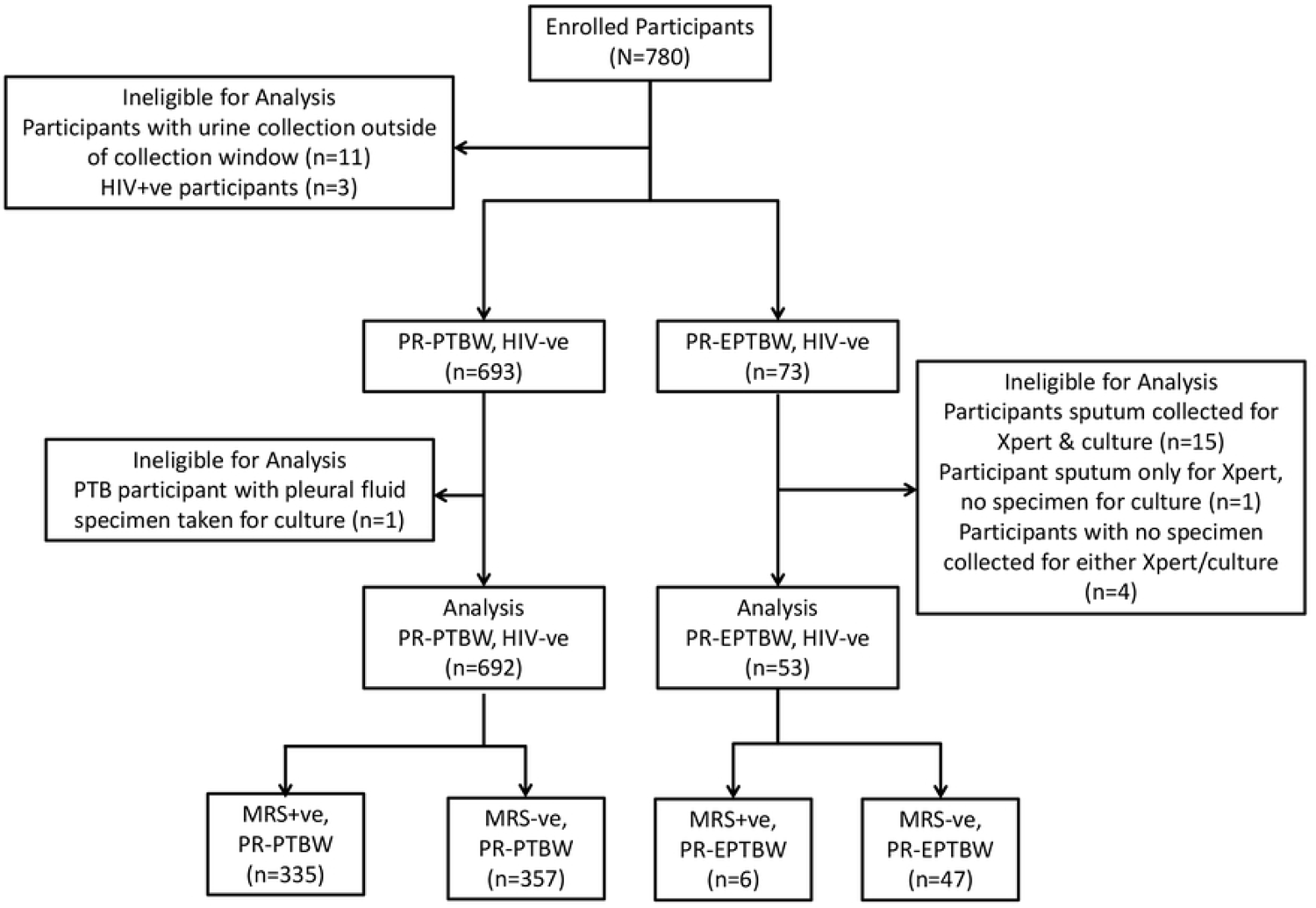
A STARD accuracy flowchart of the recruitment and diagnostic classification of the study subjects from October 2021 to April 2022. HIV, human immunodeficiency virus; PR-PTBW, PResumed Pulmonary TuBerculosis from the Wards; PR-EPTBW, PResumed Extra Pulmonary TuBerculosis from the Wards; Xpert, GeneXpert MTB/RIF Ultra.

Overall, the participants were predominantly male (66.7%), older (mean= 46.7 years, SD=16.0) and 6% self-reported a history of recent TB exposure (Table 1). The most commonly reported symptoms among the PR-PTBW group included prolonged cough (88.9%), weight loss (64.5%) and fatigue (67.6%), with fewer participants reporting prolonged fever (23.0%). High proportions of participants in the PR-EPTBW group reported typical presumptive TB symptoms (prolonged cough, fever, nights sweats, weight loss and fatigue). Chest X-ray (CXR) indicative of TB disease was seen in 21.8% of PR-PTBW with chest X-rays and 7.5% of PR-EPTBW. The majority (81.1%) of PR-PTBW were AFB smear negative, and among smear positive, nearly half had a bacterial load graded as scanty or 1+. Positivity by MGIT liquid culture was 40.5% in the PR-PTBW group and only 5.7% in the PR-EPTBW group. Xpert Ultra positivity was 43.3% in PR-PTBW, and 22.7% (5/22) in the subset of PR-EPTBW with Xpert Ultra. Overall, 335 (48.5%) of PR-PTBW and just 6 (11.3%) PR-EPTBW were microbiologically confirmed as TB positive. Using the CRS, 414 (59.8%) of PR-PTBW and 33 (62.3%) of PR-EPTBW were considered TB positive. MGIT also identified 35 participants as having an infection with nontuberculous mycobacteria (NTM).

**Table 1.** Characteristics of enrolled HIV negative participants with presumptive pulmonary and extrapulmonary TB (inpatient and outpatient recruitment, N= 745). * Captured by question “Currently living with someone with confirmed TB diagnosis?”; **496 participants with available CXR results (444 PR-PTBW, 52 PR-EPTBW); ***22 PR-EPTBW participants with non-sputum samples; denominator for PR-PTBW cohort is participants with a pulmonary specimen collected for testing, while for PR-EPTBW group is participants with a non-pulmonary specimen collected; ****Participants with inadequate specimen, NTM or not ordered set to missing for culture only reference standard.

|  | Presumptive pulmonary TB Patients (n=692) | Presumptive Extrapulmonary TB Patients (n=53) | Total (N=745) |
| --- | --- | --- | --- |
| Demographics |  |  |  |
| Female Sex, n (%) | 231 (33.4) | 17 (32.1) | 248 (33.3) |
| Age, mean (SD) | 46.6 (15.9) | 48 (17.2) | 46.7 (16.0) |
| Hospitalization status, n (%) |  |  |  |
| Inpatient | 178 (25.7) | 53 (100) | 231 (31.0) |
| Outpatient | 514 (74.3) | 0 (0) | 514 (69.0) |
| Symptom presentation, n (%) |  |  |  |
| Cough | 615 (88.9) | 44 (83) | 659 (88.5) |
| Fever | 159 (23.0) | 43 (81.1) | 202 (27.1) |
| Night sweats | 159 (23.0) | 42 (79.2) | 201 (27.0) |
| Weight Loss | 446 (64.5) | 52 (98.1) | 498 (66.8) |
| Fatigue | 468 (67.6) | 53 (100) | 521 (69.9) |
| History of recent TB exposure*, n (%) |  |  |  |
| Yes | 43 (6.2) | 2 (3.8) | 45 (6.0) |
| No | 641 (92.6) | 51 (96.2) | 692 (92.9) |
| Don't Know | 8 (1.2) | 0 (0) | 8 (1.1) |
| Chest X-Ray Indicative of TB**, n (%) | 97 (21.8) | 4 (7.5) | 101 (20.3) |
| AFB smear positive, n (%) | 131 (18.9) | 0 (0) | 131 (17.6) |
| AFB smear positivity, n (%) |  |  |  |
| Scanty | 12 (9.1) | 0 (0) | 12 (9.1) |
| 1+ | 65 (49.6) | 0 (0) | 65 (49.6) |
| 2+ | 32 (24.4) | 0 (0) | 32 (24.4) |
| 3+ | 22 (16.8) | 0 (0) | 22 (16.8) |
| Xpert Ultra positive***, n (%) | 300 (43.3) | 5 (22.7) | 305 (42.7) |
| Rif resistance detected, n (%) | 12 (4.0) | 0 (0) | 12 (1.6) |
| MGIT liquid culture positivity, n (%) |  |  |  |
| Positive | 280 (40.5) | 3 (5.7) | 283 (37.9) |
| NTM | 35 (5.1) | 0 (0) | 35 (4.7) |
| Negative | 375 (54.2) | 50 (94.3) | 425 (50.3) |
| Specimen contamination/not ordered | 2 (0.3) | 0 (0) | 2 (0.3) |
| Microbiological Reference Standard, n (%) | 335 (48.5) | 6 (11.3) | 341 (45.8) |
| Xpert Ultra positive***, n (%) | 300 (43.4) | 5 (22.7) | 305 (42.7) |
| Culture positive****, n (%) | 280 (42.7) | 3 (5.7) | 283 (40.0) |
| Clinical Reference Standard, n (%) | 414 (59.8) | 33 (62.3) | 447 (60.0) |

The analysis of the uLAM concentrations showed the FIND 28:A194-01 (F/A) Ab pair had a greater number of positive tests in both PR-PTBW and PR-EPTBW cohorts (Table 2). A similar percentile of PR-PTBW and PR-EPTBW had uLAM were detected via F/A (30.6% vs 34.0%, *p*=0.643) and also S4-20/A194-01 (S/A; 20.1% vs 15.1%, *p*=0.475) when not stratifying by MRS status. When pooled, F/A detected positivity was 230 (30.9%) while S/A was 147 (19.7%). Among participants with uLAM detected by F/A, the median estimated uLAM concentration was similar with PR-PTBW at 79.0 pg/mL (interquartile range [IQR] 31.8 - 290.5), and 76.5 pg/mL (IQR 34.3-190.0) for PR-EPTBW. When stratifying by MRS status, median estimated uLAM concentration appeared higher for MRS-positive PR-PTBW (110.0 pg/mL vs. 45.0 pg/mL) compared to MRS-negative PR-PTBW, higher among MRS-negative compared to MRS-positive PR-EPTBW (44.0 pg/mL vs 77.0 mg/mL). Among participants with uLAM detected in their urine using the S/A pair, the median estimated uLAM concentration among PR-PTBW was 55.0 pg/mL, (IQR 27.0 - 139.5), and 36.0 pg/mL (IQR 29.0-46.3) for PR-EPTBW. When stratifying by MRS status, median estimated uLAM concentration appeared higher for MRS-positive PR-PTBW (57.0 pg/mL vs. 21.0 pg/mL) compared to MRS-negative PR-PTBW, as well as for PR-EPTBW (50.5 pg/mL vs. 33.0 pg/mL) respectively. When viewing the distribution of estimated uLAM concentrations for the same participants using the two antibody pairs, the F/A pair appears to estimate a higher concentration of 78.0 pg/mL as compared to the S4-20:A194-01 Ab pair with 51.0 pg/mL (Table 2 and Figure 2). Only 87 (32.9%) PR-PTBW and 5 (23.8%) PR-EPTBW were uLAM positive by both F/A and S/A pairs, indicating a high degree of discordant detection (S1 Fig).

**Figure 2.**
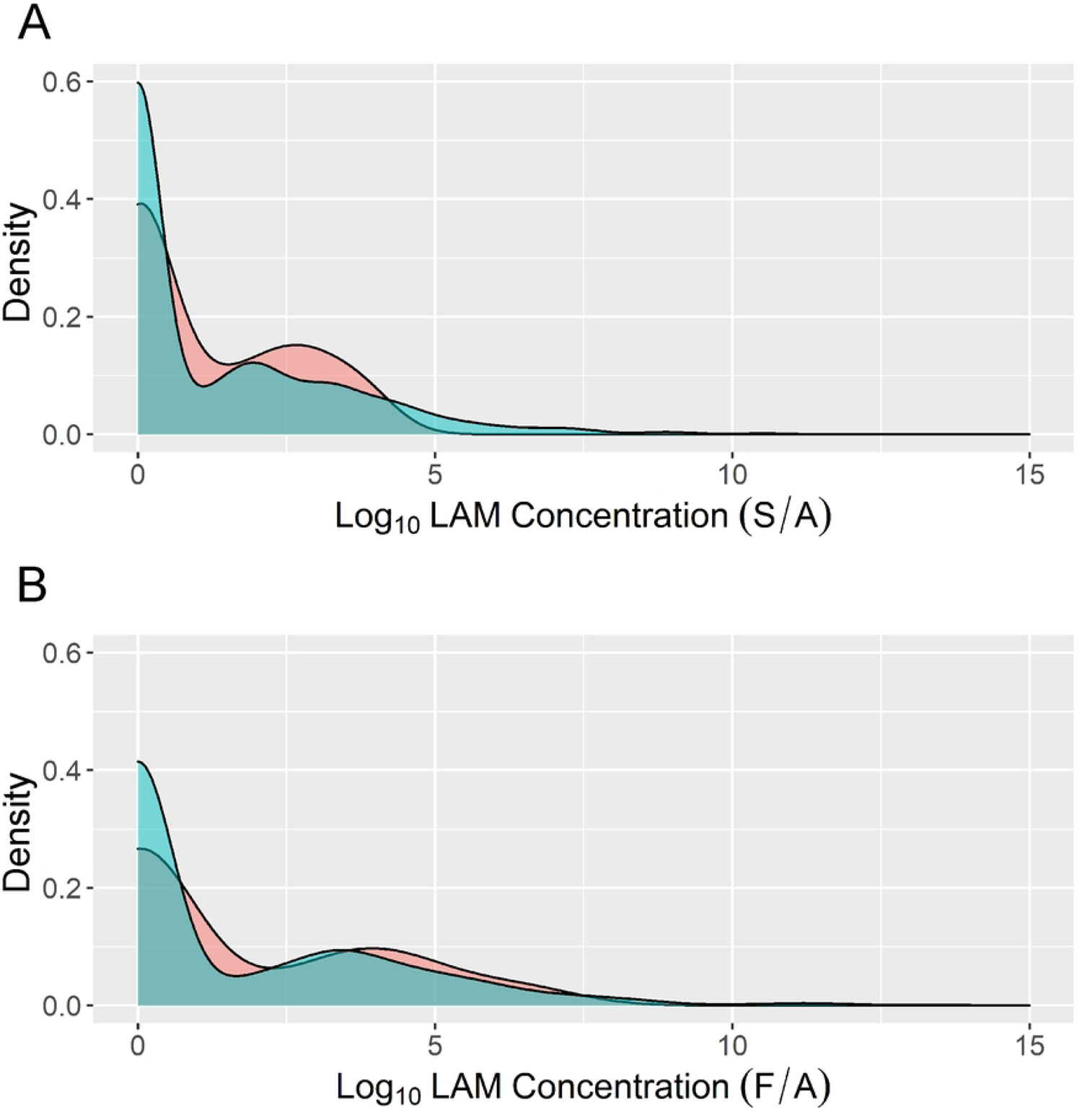
The range of uLAM concentrations quantitated by the ECL immunoassay using antibody pairs S/A and F/A (N=745).

**Table 2.** ECL test results of study participants using S/A and F/A Ab pairs (N=745). The number of S/A and F/A positive results for both PR-PTBW and PR-EPTBW, and the pooled data with the median uLAM concentration (pg/mL) among participants with detected uLAM. Abbreviations: PR-PTBW, presumptive pulmonary TB from ward; PR-EPTBW, presumptive extra pulmonary TB from ward; MRS, microbiological reference standard; ECL, electrochemiluminescent; S/A, S4-20/A194-01 Ab pair; N, number; F/A, FIND28/A194-01 Ab pair. IQR, interquartile range.

| ECL Test Results | PR-PTBW (N=692) |  | PR-EPTBW (N=53) |  | Pooled (N=745) |
| --- | --- | --- | --- | --- | --- |
|  | MRS + (n=335) | MRS - (n=357) | MRS + (n=6) | MRS - (n=47) |  |
| S/A positive result – N (%) | 130 (38.8) | 9 (2.5) | 2 (33.3) | 6 (12.8) | 147 (19.7) |
| S/A Median uLAM conc. – pg/mL (IQR) | 57.0 (29.0-150.0) | 21.0 (17.0 – 26.0) | 50.5 (50.25 – 50.75) | 33.0 (23.0 – 37.0) | 51.0 (27.0 – 125.0) |
| F/A Positive result – N (%) | 137 (40.9) | 75 (21.0) | 3 (50.0) | 15 (31.9) | 230 (30.9) |
| F/A Median uLAM conc. – pg/mL (IQR) | 110.0 (36.0-374.0) | 45.0 (29.5 – 104.0) | 44.0 (30.5-93.0) | 33.0 (23.0 – 37.0) | 78.0 (31.2 – 284.0) |

When evaluating the diagnostic performance of uLAM detection using the S/A pair the overall senstivity was low in this HIV negative population. A similar proportion of microbiologically confirmed participants were correctly identified in PR-PTBW (sensitivity: 39%, 95% CI: 34-44%) and PR-EPTBW (sensitivity: 33%, 95% CI: 4-78%), (Table 3). However, true negatives were more accurately identified in PR-PTBW (specificity: 97%, 95% CI: 95-99%) compared to PR-EPTBW (specificity: 87%, 95% CI: 74-95%). When using the CRS as the reference standard instead, test sensitivity was 32% (95% CI: 28-37%) in PR-PTBW and 24% (95% CI: 11 – 42%) in the PR-EPTBW, and specificity was very similar in both groups at 98%-100%. When the immunoassay data for S/A and F/A was pooled and also compared to the MRS, Xpert Ultra, MGIT and CRS data for both PR-PTBW (S2 Table) and PR-EPTBW (S3 Table). The sensitivity for PR-PTBW was 55% (95% CI: 0.490.60%) and the specificity was 78% (95% CI: 0.73-0.82%). With the PR-EPTBW group, sensitivity was similar at 50% (95% CI: 0.12-0.88%) and with a specificity of 62% (95% CI: 0.46-0.75%).

**Table 3.** The diagnostic sensitivity and specificity of the ECL S/A and & F/A immunoassays in the PR-PTBW and PR-EPTBW groups as compared to the microbiologic reference standards (MRS), Xpert, MGIT, and the clinical reference standard (CRS) used in the diagnosis of TB disease. The positive predictive values (PPV), negative predictive values (NPV), positive likelihood ratios (PLR) and negative likelihood ratios (NLR) were also calculated for each. Abbreviations: N, number; TP, true positive; TN, true negative; FN, false negative; FP, false positive; CI, confidence interval; S/A, S4-20/A194-01; F/A, FIND28/A194-01; PR-PTBW, presumed pulmonary tuberculosis from ward; MGIT, mycobacterial growth indicator tube; MRS, microbiological reference standard; CRS, clinical reference standard; PR-EPTBW, presumed extra pulmonary tuberculosis from ward.

|  |  | Positive by reference standard |  | Negative by reference standard |  |  |  |  |  |
| --- | --- | --- | --- | --- | --- | --- | --- | --- | --- |
| Reference Standard | N | TP/TP+FN | Sensitivity (95% CI) | TN/TN+FP | Specificity (95% CI) | PPV (95% CI) | NPV (95% CI) | PLR (95% CI) | NLR (95% CI) |
| S/A PR-PTBW |  |  |  |  |  |  |  |  |  |
| MRS | 692 | 130/335 | 0.39 (0.34, 0.44) | 348/357 | 0.97 (0.95, 0.99) | 0.94 (0.88, 0.97) | 0.63 (0.59, 0.67) | 15.4 (7.96, 29.75) | 0.63 (0.58, 0.68) |
| Xpert Ultra | 692 | 123/300 | 0.41 (0.35, 0.47) | 376/392 | 0.96 (0.93, 0.98) | 0.88 (0.82, 0.93) | 0.68 (0.64, 0.72) | 15.4 (7.96, 29.75) | 0.62 (0.56, 0.68) |
| MGIT<sup>§</sup> | 655 | 112/280 | 0.40 (0.34, 0.46) | 356/375 | 0.95 (0.92, 0.97) | 0.95 (0.92, 0.97) | 0.68 (0.64, 0.72) | 7.89 (4.98, 12.52) | 0.63 (0.57, 0.70) |
| CRS | 692 | 133/414 | 0.32 (0.28, 0.37) | 272/278 | 0.98 (0.95, 0.99) | 0.96 (0.91, 0.98) | 0.49 (0.45, 0.53) | 14.88 (6.66, 33.25) | 0.69 (0.65, 0.74) |
| S/A PR-EPTBW |  |  |  |  |  |  |  |  |  |
| MRS | 53 | 2/6 | 0.33 (0.04, 0.78) | 41/47 | 0.87 (0.74, 0.95) | 0.25 (0.03, 0.65) | 0.91 (0.79, 0.98) | 2.61 (0.67, 10.13) | 0.76 (0.43, 1.36) |
| Xpert Ultra* | 22 | 2/5 | 0.40 (0.05, 0.85) | 11/17 | 0.65 (0.38, 0.86) | 0.25 (0.03, 0.65) | 0.79 (0.49, 0.95) | 1.13 (0.32, 3.96) | 0.93 (0.42, 2.06) |
| MGIT | 53 | 1/3 | 0.33 (0.01, 0.91) | 43/50 | 0.86 (0.73, 0.94) | 0.12 (0, 0.53) | 0.96 (0.85, 0.99) | 2.38 (0.42, 13.59) | 0.78 (0.35, 1.74) |
| CRS | 53 | 8/33 | 0.24 (0.11, 0.42)) | 20/20 | 1.00 (0.83, 1.00) | 1.00 (0.63, 1.00) | 0.44 (0.30, 0.60) | Inf (NA, Inf) | 0.76 (0.62, 0.92) |
| F/A PR-PTBW |  |  |  |  |  |  |  |  |  |
| MRS | 692 | 137/335 | 0.41 (0.36, 0.46) | 282/357 | 0.79 (0.74, 0.83) | 0.65 (0.58, 0.71) | 0.59 (0.54, 0.63) | 1.95 (1.53, 2.47) | 0.75 (0.67, 0.83) |
| Xpert Ultra | 692 | 124/300 | 0.41 (0.36, 0.47) | 304/392 | 0.78 (0.73, 0.82) | 0.58 (0.52, 0.65) | 0.63 (0.59, 0.68) | 1.84 (1.47, 2.31) | 0.76 (0.68, 0.84) |
| MGIT<sup>§</sup> | 655 | 118/280 | 0.42 (0.36, 0.48) | 290/375 | 0.77 (0.73, 0.81) | 0.58 (0.51, 0.65) | 0.64 (0.60, 0.69) | 1.86 (1.47, 2.34) | 0.75 (0.67, 0.84) |
| CRS | 692 | 154/414 | 0.37 (0.33, 0.42) | 220/278 | 0.79 (0.74, 0.84) | 0.73 (0.66, 0.79) | 0.46 (0.41, 0.50) | 1.78 (1.37, 2.31) | 0.79 (0.72, 0.87)v |
| F/A PR-EPTBW |  |  |  |  |  |  |  |  |  |
| MRS | 53 | 3/6 | 0.50 (0.12, 0.88) | 32/47 | 0.68 (0.53, 0.81) | 0.17 (0.04, 0.41) | 0.91 (0.77, 0.98) | 1.57 (0.64, 3.86) | 0.73 (0.32, 1.67) |
| Xpert Ultra* | 22 | 2/5 | 0.40 (0.05, 0.85) | 11/17 | 0.65 (0.38, 0.86) | 0.25 (0.03, 0.65) | 0.79 (0.49, 0.95) | 1.13 (0.32, 3.96) | 0.93 (0.42, 2.06) |
| MGIT<sup>§</sup> | 53 | 2/3 | 0.67 (0.09, 0.99) | 34/50 | 0.68 (0.53, 0.80) | 0.11 (0.01, 0.35) | 0.97 (0.85, 1) | 2.08 (0.85, 5.11) | 0.49 (0.10, 2.46) |
| CRS | 53 | 13/33 | 0.39 (0.23, 0.58) | 15/20 | 0.75 (0.51, 0.91) | 0.72 (0.47, 0.90) | 0.43 (0.26, 0.61) | 1.58 (0.66, 3.76) | 0.81 (0.56, 1.17) |

<sup>§</sup> Participants with inadequate specimen, NTM, or not ordered set to missing for culture only reference standard.
\* 22 participants had a non-sputum sample taken for Xpert Ultra testing for EPTB; participants with a sputum sample taken were excluded from analysis. The 22 shown due to having a non-sputum sample for Xpert Ultra testing are likely not a random sample of the total 53 participants in the table, and diagnostic performance estimates should be interpreted with caution.

When evaluating the diagnostic performance of uLAM detection using the F/A pair, sensitivity was 41% (95% CI: 36-46%) in PR-PTBW, and 50% (95% CI: 12-88%) in PR-EPTBW group (*p*=0.693) (Table 3). True negatives may have been somewhat more accurately identified using F/A in PR-PTBW (Specificity: 79%, 95% CI: 74-83%) compared to PR-EPTBW (Specificity: 68%, 95% CI: 53-81%, *p*=0.096). When comparing performance using the CRS as the reference standard, F/A test sensitivity and specificity were very similar in PR-PTBW and PR-EPTBW, with sensitivity of 37%-39% and specificity of 75-79%.

When assessing performance between the S/A and F/A Ab pairs, their sensitivity was similar in the PR-PTBW (*p*=0.356). However, the S/A pair had significantly higher specificity (97% vs 79%, *p<0.0001*) and PPV (94% vs 65%, *p<0.0001*). Similar diagnostic performances were observed between both Ab pairs in PR-PTBW when using CRS as the reference standard. Amongst PR-EPTBW, while sensitivity against MRS appeared to be slightly higher with F/A, a detectable difference in performance was not observed in this small EPTB MRS positive group (n=6, 50% vs 33%, *p*>0.99). However, specificity was higher for the S/A than for F/A pair (87% vs 68%, *p*=0.039) in PR-EPTBW. When comparing the discriminatory ability at varying estimated concentration thresholds, the S/A pair showed greater discriminatory ability compared to F/A pair in PR-PTBW (area under the curve [AUC]=0.744 vs 0.628, *p*<0.0001) and PR-EPTBW (AUC=0.755 vs 0.5339, *p*=0.045) (Figure 3).

**Figure 3.**
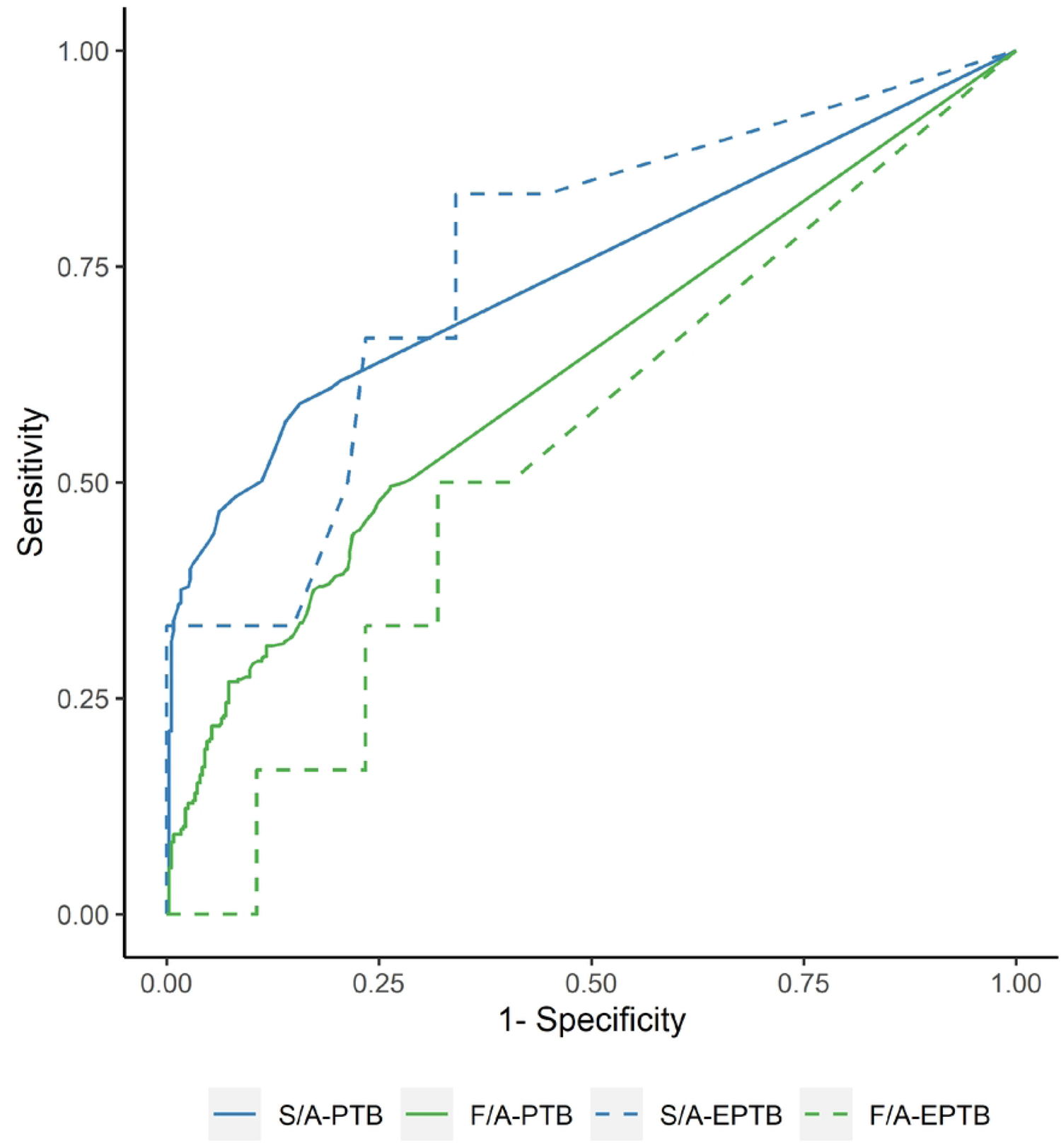
Receiver operating characteristic curves comparing estimated uLAM concentrations (pg/mL) derived from the S/A and F/A Ab pairs to microbiological reference standard (MRS), all groups.

Receiver operating curves comparing the ECL immunoassay uLAM concentrations to the MRS, using 2 separate antibody pairs for both PR-PTBW & PR-EPTBW. Comparison to the MRS using the S4-20:A194-01 (S/A) pair is shown in blue, with performance PR-PTBW (AUC=0.744, n=692) shown in the solid line and performance in PR-EPTBW (AUC=0.755, n=53) shown in the dashed line. Comparison to the MRS using the FIND 28:A194-01 (F/A) pair is shown in green, with performance in PR-PTBW (AUC, 0.628, n=692) shown in the solid line and performance in PR-EPTBW (AUC=0.539, n=53) shown in the dashed line. Discriminatory ability using the S4-20:A194-01 Ab pair was high for both PR-PTBW & PR-EPTBW compared to the FIND-28:A194-01 pair.

## Discussion

We have completed the largest study to date on a HIV negative cohort presenting with presumptive TB disease wherein we assessed the diagnostic performance of a highly sensitive immunoassay platform to detect the levels of uLAM from urine as compared to the MRS. In a cohort of HIV-uninfected adults with either presumptive PTB or EPTB disease, uLAM was detectable in less than half of people and the concentrations observed were typically low. The F/A pair had slightly higher sensitivity than the S/A pair (41% vs. 39%) but also had a much lower specificity (79% vs. 97%). The routine standard of care included sputum smear microscopy wherein the majority of PTB patients were sputum smear-negative (18.9% smear positivity), suggesting that a large proportion of participants in this study had subclinical TB with a relatively low bacterial disease burden and thus less availability of LAM at the site of infection and potentially also in the urine. Others have also noted significant differences in uLAM levels based on the geographical sites, presumably highlighting that different TB lineages may affect the levels of uLAM observed in patient urine [15,19]. We also investigated the performance of the immunoassays with urine collected from either pulmonary or EPTB. No significant differences was seen between these groups with either antibody pair although it should be noted the size of the EPTB group (N = 53, MRS TB positive 6) was much smaller than the PTB group (N = 692, MRS TB positive 335).

Many previous cohorts that have been screened for uLAM in urine typically have a higher prevalence of HIV positive comorbidity and as such often have higher concentrations of uLAM in their urine. The results from this study are less optimistic than from other studies who reported significantly higher sensitivities when detecting uLAM in HIV negative cohorts [24–26]. Broger et al analyzed concentrated urine from 372 HIV-negative participants from Peru and South Africa using the S/A assay on the MSD platform with the pooled performance data having a sensitivity of 66.7% (95% CI 57.5%-74.7%) and a specificity of 98.1% (95% CI 95.6%-99.2%) [19]. It was also noted in the supplemental data in a subgroup analysis that the sensitivity was much higher in Peru (78.5%) compared with South Africa (37.5%), with specificities of 100% and 95.3% respectively, the latter data reflecting that the poor sensitivity of uLAM detection on the MSD platform has been observed [19]. While we used the same platform and ECL assays as previously used in an earlier smaller scale study on an HIV negative cohort [20], we used a different methodology for establishing the cut off. In the work by Sigal et al. they assigned cut off value of 11 pg/mL to the S/A assay results [20], while Broger et al used a cutoff of 5.4 pg/mL [19], noting that such a cutoff would not be applicable to other LAM assays. Where a cutoff allows for practicality and comparative analysis, using this ultrasensitive method for uLAM is still relatively new. Furthermore, using plate based LOD allows for a more standardized and objective measure of a given analyte, especially one as heterogenous as LAM [9,15,25], where there are inherent differences in the antibody pairs being assessed respective to sensitivity and specificity. Once use of a specific antibody pair is normalized, a generalizable cutoff would be expeditious in studying LAM in various populations. The negative association of glycosuria on uLAM concentration with A194-01 and an S4-20 derivative has been noted, we did not perform urinalysis on the samples used and so a greater proportion of participants with glycosuria would influence our results [25].

Other studies have measured uLAM using different methods. One study reported the use of nanocages hosting a dye as the capture ligand in association with three detector antibodies including A194-01 and MoAB1, a recombinant derivative of S4-20 [27]. The nanocages offered a 50-fold increase in uLAM concentration and overall, the pooled diagnostic accuracy from the three antibodies was 90% sensitivity at 73.5% specificity [25,27]. A further study had 160 participants who were predominantly HIV negative and this study also reported very high sensitivity and specificity using the antibody pair CS-35/A194-01 but the authors acknowledge that the study focused on predominantly TB positive cohorts (140 TB positive from 160 samples, many of which were smear 1+ or greater) and so diagnostic accuracy was not established due to the low number of TB negative cases included (N = 10) [28]. This study also included proteinase K pretreatment and mild heating (55°C) which significantly improved the sensitivities of the immunoassay used. In addition, all samples were derived for Peru and we speculate that this data augments the observations for Broger et al where significantly more uLAM was seen in samples from this country.

There are several limitations to this study. First, all of the ECL testing was conducted on previously frozen and not fresh urine specimens. While banked samples are more convenient for high throughput testing, long-term freezing may diminish uLAM levels [29,30]. While samples were maintained at −80°C on the day of collection until the time of testing, we did not assess the time from specimen collection to freezing. The clinical laboratory protocol did note to store the samples at 4°C soon after collection and to then store aliquots at −80°C once 15 mL conical tubes had been filled. There may also have been additive effects caused by the COVID-19 pandemic on exposure risk to TB while under lock down and in changes to care seeking behaviors due to the pandemic. The powered calculations for the study protocol were based on pre-pandemic national TB prevalence rates wherein an estimated 271 confirmed TB positive cases would be identified within a cohort of 780. Further, greater awareness of the risks of respiratory disease may have induced earlier care seeking in some cases and as such cases of TB were identified earlier and in addition lack of access to care during lock downs may have led to cases of self-clearance of TB.

The predominant lineage of the *Mycobacterium tuberculosis* strain may also play a role in variation of the median level of uLAM detected [15,25]. In this work, we also took a slightly different approach to ascribing a detection threshold to score the presence of uLAM based on the LOD determined on a plate-by-plate basis via each standard curve versus a uniform cut off for all data [19–20]. While we have confidence in the quality of the data generated to describe the PTB cohort based on the number of participants and the relative ease with which diagnostic results could be applied to score as PTB or TB negative. The study was not designed to enroll a sufficient number of PR-EPTBW to allow statistical comparisons between PR-EPTBW and PR-PTBW; 53 versus 692 respectively and added to this were limitations in the collection and use of appropriate samples to microbiologically confirm EPTB diagnosis. Finally, the prevalence of bacteriologically confirmed TB is higher than expected in a population of presumptive adults presenting at both outpatient and inpatient settings. It is important to note that study enrollment coincided with Vietnam’s surge in SARS-CoV-2 infections associated with the spread of the Omicron variant, which could have potentially affected healthcare-seeking behavior due to mobility restrictions and increased screening at health facilities. This could potentially explain the low proportion of reported febrile illness observed in this cohort, and could potentially result in patients only seeking care with increased TB disease severity. However, the higher proportion of AFB smear microscopy results with scanty/1+ grading indicate that this may not be the case. Nonetheless, it is important to take this into consideration when evaluating the positive and negative predictive test values, which are influenced by underlying disease prevalence.

Biomolecules in urine samples may present challenges to sensitivity, and several studies have investigated specimen preparation methods to improve access for the antibody to its target epitope [31]. Improved methods include specimen preparation via enzymatic or inhibitor specific treatments to release sequestered uLAM from complexes or remove inhibitory compounds such as lipids or protein-uLAM complexes [32–34]. The enrichment of uLAM concentration in urine specimens has been achieved via a variety of methods using either the direct capture of uLAM via specific antibodies [35,36], a uLAM specific chemical ligand or via proteolysis, chemical treatment and ultrafiltration [27,37]. Others have focused on improving high sensitivity assays via platform improvement, novel antibodies, and the application of machine learning algorithms to improve diagnostic accuracy [38–40]. The application of multiple antibodies, each targeting different LAM epitopes, has also been demonstrated to increase diagnostic performance [25]. A key component to this work is to ensure that clinical samples are used early in the product development phase and that clinical evaluation should encompass multiple populations as evidenced in this and other work where the detection of uLAM in the urine of TB positive HIV negative patients varies [19,28,38].

In conclusion, from a large HIV-negative cohort in Vietnam, high-sensitivity immunoassay testing for uLAM did not reach the levels noted in the TPP criteria for an effective diagnostic test for TB disease. Developing a better uLAM assay to diagnose the majority of people with TB disease may require novel antibodies, a urine concentration step, or both. Nonetheless, developing non-sputum based RDTs to diagnose people with TB disease needs to remain a global health priority.

## Data Availability

All of the data generated in this study can be publicly accessed at Dataverse (https://doi.org/10.7910/DVN/AOL0LP). This includes all of the relevant clinical and statistical data and also the data from the laboratory testing.

## Acknowledgements

The project team would like to thank each of the participants in this study for their willingness to enroll, provide urine samples to support this work, and to permit the use of their test data. We would also like to acknowledge the staff of the National Lung Hospital of Hanoi for their dedication and commitment to the implementation and operation of this project.

## Supplemental Materials

**S1 Table.**
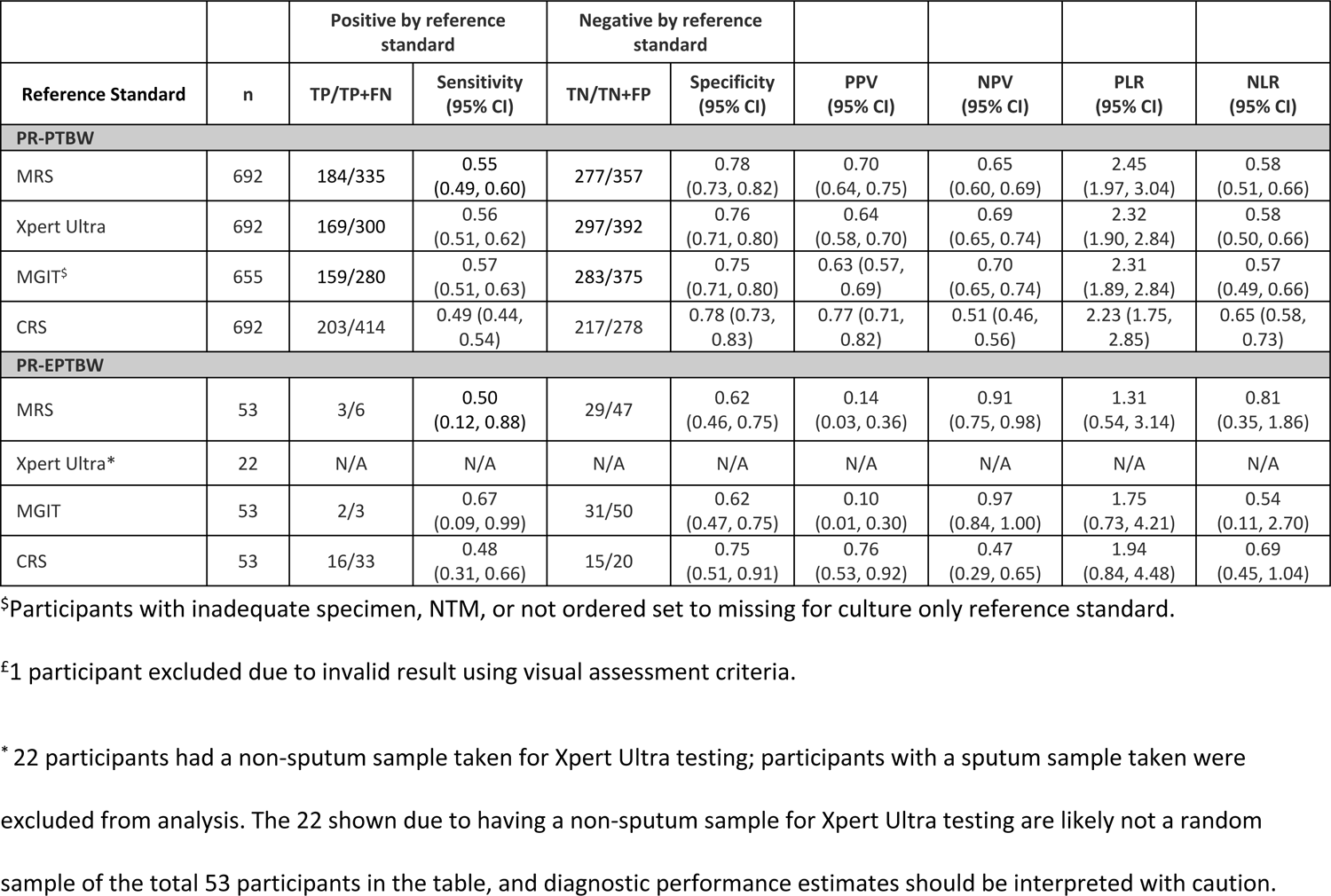
The diagnostic sensitivity and specificity of the pooled ECL immunoassays antibody pairs. The PR-PTBW and PR-EPTBW groups compared to the MRS, Xpert, MGIT and the CRS were used for the diagnosis of active TB. Abbreviations: N, number; TP, true positive; TN, true negative; FN, false negative; FP, false positive; CI, confidence interval; PPV, positive predictive value; NPV, negative predictive value; PLR, positive likelihood ratio; NLR, negative likelihood ratio S/A, S4-20/A194-01; F/A, FIND28/A194-01; PR-PTBW, presumed pulmonary tuberculosis from ward; MGIT, mycobacterial growth indicator tube; MRS, microbiological reference standard; CRS, clinical reference standard; PR-EPTBW, presumed extra pulmonary tuberculosis from ward; N/A, not applicable.

**S2 Table.** Plate lower limits of detection (LLOD) of PR-PTBW participants. The LLODs from the immunoassay plates used for uLAM quantitation using the S4-20/A194-01 immunoassay stratified by MRS status from the PR-PTBW group (n=692). Abbreviations: N, number; MRS, microbiological reference standard; N/A, not applicable.

| LLOD<br>(pg/mL) | n (%) | uLAM Detected (n=139) |  | uLAM Not Detected (n=553) |  |
| --- | --- | --- | --- | --- | --- |
|  |  | Mean (min, max) | n | Mean (min, max) | n |
| 10 | 16 | 129.5 (22, 346) | 4 | 0.1 (0,1) | 12 |
| 11 | 111 | 162.0 (12, 2160) | 27 | 1.1 (0, 8) | 84 |
| 12 | 39 | 902.8 (17, 7039) | 10 | 1.1 (0, 10) | 29 |
| 13 | 79 | 224.0 (31, 599) | 8 | 1.5 (0, 8) | 71 |
| 15 | 57 | 216.9 (15, 1691) | 13 | 1.2 (0, 10) | 44 |
| 16 | 34 | 197.2 (17, 2138) | 15 | 3.3 (0, 12) | 19 |
| 17 | 39 | 1582.7 (21, 9244) | 6 | 1.6 (0, 11) | 33 |
| 18 | 117 | 404.0 (18, 6017) | 24 | 2.1 (0, 16) | 93 |
| 19 | 37 | 12443.3 (41, 36994) | 3 | 2.4 (0, 14) | 34 |
| 21 | 75 | 82.2 (21, 299) | 16 | 1.8 (0, 16) | 59 |
| 22 | 8 | 32.0 (32, 32) | 1 | 1.9 (0, 11) | 7 |
| 23 | 38 | 653.5 (71, 1236) | 2 | 2.5 (0, 14) | 36 |
| 26 | 39 | 369.6 (27, 1235) | 7 | 3.3 (0, 18) | 32 |
| 52 | 1 | 69.0 (69, 69) | 1 | NA | 0 |
| 63 | 2 | 94.5 (68, 121) | 2 | NA | 0 |

**S3 Table.** Plate lower limits of detection (LLOD) of PR-EPTBW participants. The LLOD from the immunoassay plates used for uLAM quantitation using the S4-20/A194-01 immunoassay stratified by MRS status from the PR-EPTBW group (N=53). Abbreviations: N, number; MRS, microbiological reference standard; N/A, not applicable.

| LLOD<br>(pg/mL) | n (%) | LAM Detected (n=8) |  |  |  | LAM Not Detected (n=45) |  |  |  |
| --- | --- | --- | --- | --- | --- | --- | --- | --- | --- |
|  |  | MRS Positive<br>(n=2) |  | MRS Negative<br>(n=6) |  | MRS Positive<br>(n=2) |  | MRS Negative<br>(n=6) |  |
|  |  | Mean<br>(min, max) | n | Mean<br>(min, max) | n | Mean<br>(min, max) | n | Mean<br>(min, max) | n |
| 10 | 4 (7.5) | - | 0 | - | 0 | - | 0 | 0.5 (0, 2) | 4 |
| 13 | 1 (1.9) | - | 0 | - | 0 | 10.0 (10, 10) | 1 | - | 0 |
| 16 | 4 (7.5) | 51.0 (51, 51) | 1 | 24.0 (18, 34) | 3 | - | 0 | - | 0 |
| 22 | 26 (49.1) | 50.0 (50, 50) | 1 | - | 0 | 4.0 (4, 4) | 1 | 1.9 (0, 15) | 24 |
| 27 | 18 (34) | - | 0 | 38.3 (32, 45) | 3 | 7.5 (0, 15) | 2 | 5.5 (0, 21) | 13 |

**S1 Fig.** Urine Log LAM concentration by the ECL immunoassay using antibody pairs S/A and F/A (N=745). A comparison of the distribution of estimated uLAM concentrations for the same participants using S/A and F/A the two antibody pairs.

**S1 Doc.** Participant inclusion and exclusion criteria.

**S2 Doc.** Study Procedures

